# Direct sequencing of *Leishmania donovani* from patients in Garissa County, Northern Kenya, reveals a newly emerging intra-specific hybrid genotype

**DOI:** 10.1101/2025.05.15.25327683

**Authors:** Vane Kwamboka Omwenga, Kimita Gathii, Cyrus Ayieko, Clement Masakhwe, Mohamed Hussein Ibrahim, Senne Heeren, Jean Claude Dujardin, Pieter Monsieurs, Malgorzata Anna Domagalska, John N Waitumbi

## Abstract

**Introduction:** Leishmaniasis is endemic in many countries, including Kenya. Despite the rising frequency of visceral leishmaniasis (VL) outbreaks in Garissa County, Northeastern Kenya, there is limited data on the genetic structure and epidemiology of *Leishmania* parasites from this region. This study used molecular methods to characterize *Leishmania* parasites collected at Garissa County Referral Hospital during the 2019 - 2022 VL outbreak.

**Methods:** 286 blood samples, collected from patients suspected of having VL at Garissa County Referral Hospital between 2019 and 2022 were used. *Leishmania* parasites were screened at genus level by a quantitative real-time PCR assay targeting the arginine permease gene AAP3 (AAP3-qRT-PCR). Species identification and targeted gene sequencing were made on Illumina MiSeq using PCR amplicons of *Hsp70* gene and ITS regions. Whole genome sequencing (WGS) was performed directly on eight selected blood samples using a target enrichment method after which data was analyzed using phylogenomic tools.

**Results:** By AAP3-qRT-PCR, 128/286 (45%) blood specimens were determined to have *Leishmania* parasites. We obtained 86 *Hsp70* and 79 ITS sequences that phylogenetically clustered with the *L. donovani* species complex. By WGS, the eight selected samples had *L. donovani* s.s., and clustered in two separate groups: one similar to the previously reported *L. donovani* group 5 and the other constituted a new and intra-specific hybrid genetic variant not reported previously. In all the 8 Kenya samples, we found SNPs in genes previously shown to be involved in *L. donovani* resistance to Antimony, Amphotericine B and Miltefosine.

**Conclusion:** This study reveals the complex nature of *Leishmania* genetic structure in Kenya and sheds light on the genomic polymorphism of *L. donovani* in the region, which in turn, may explain the evolving threat of VL in the region. As caveat, the genomic signatures of drug resistance genes that were identified should be interpreted with caution until their functional implication is clarified in future studies.

**Non-technical Summary:** Visceral leishmaniasis (VL), or kala-azar, is a deadly disease if untreated. In Kenya, it is common in Garissa, Isiolo, Marsabit, Turkana, and Wajir. During the 2019-2022 outbreak, we collected samples from patients at Garissa County Referral Hospital to study the genetic structure of the parasites. To avoid biases from lab-grown parasites, we used advanced genetic methods on parasites directly from patients’ blood. We identified two parasite groups: one related to a known group 5 (Ld5) from Kenya and Ethiopia, and a new group with mixed ancestry from Sudan, Ethiopia (Ld3), and Iraq (Ld4). Notably, we found genes linked to drug resistance. These findings are crucial because they help in understanding how the parasites evolve and spread. As caveat, the presence of drug resistance genes should be interpreted with caution until their functional implication are clarified in future studies.

## Introduction

Leishmaniasis is a life-threatening vector-borne tropical disease caused by protozoan parasites of the genus *Leishmania* and transmitted by female Phlebotomine sand flies [1]. The disease is endemic in 99 nations including Kenya (https://www.who.int/data/gho/data/themes/topics/gho-ntd-leishmaniasis), and is responsible for roughly 1 million cases and 50,000 fatalities annually [2]. The disease presents in three main forms influenced by the infecting species [3,4]: cutaneous leishmaniasis (CL) and its subtypes; visceral leishmaniasis (VL, also known as Kala azar), [5]; and muco-cutaneous leishmaniasis (MCL) [6]. Of these, VL is the most serious and ranks as the second deadliest protozoan disease after malaria [7].

The distribution of leishmaniasis varies geographically with approximately 90% of global visceral cases occurring in Sub-Saharan Africa, Southeast Asia, and Brazil [7]. For a long time, the hotspot for VL was concentrated in the Indian subcontinent (ISC), but due to the success of recent elimination program in the ISC, East Africa now appears to have become a VL hotspot, where there is limited elimination progress [8]. Some of the key factors that contributed to near elimination of VL in ISC included political commitment, targeted implementation that included treatments and follow-up of cases, strong surveillance among others [9,10]. East Africa, leishmaniasis is widespread in Kenya, Ethiopia, Sudan, Uganda, Somalia and Eritrea [11]. Kenya specifically reports endemicity of the disease in the Rift Valley, Eastern and Northeastern regions, with outbreaks of VL occurring in sub-counties of Garissa, Isiolo, Marsabit, Turkana and Wajir [12], with approximately 5 million people being at risk of infection annually [13]. This underscores the urgency of conducting concerted epidemiological research to better understand and combat the disease in the country. The classification of *Leishmania* species remains contentious, with up to 53 species identified, 21 of which are known to infect humans [5,14]. Based on their geographical distribution, these species are grouped into New World -found in the Americas-, and Old World -distributed across Europe, Asia and Africa [15,16]. In Eastern Africa, VL is primarily attributed to two species of the *donovani* complex, *L. donovani* and *L. infantum*. Notably, the geographical distribution of these species can lead to genetic variations due to changes in virulence genes and potential hybrid formation, complicating our understanding of leishmaniasis [17–20].

This study used targeted gene sequencing and untargeted WGS to directly characterize the *Leishmania* parasites collected from patients’ blood at Garissa County Referral Hospital, in Northeastern Kenya during the 2019 - 2022 VL outbreak. Noteworthy, we applied here for the first time in Africa, a method for direct genome sequencing of *L. donovani* in blood samples from VL patient [21,22]. Garissa and neighboring counties such as Kitui experience recurring outbreaks, probably because the semi-arid ecology provides ideal conditions for sand flies, particularly around termite mounds that offer microhabitats with moisture and organic matter for breeding [23]. Additionally, poverty, limited healthcare access, overcrowding and poor housing conditions especially in refugee camps like Dadaab, intensify disease spread [24].

## Materials and Methods

### Ethics statement

This study was conducted as part of a public health response following a request from the Ministry of Health, Garissa County to support a *Leishmania* outbreak investigation. WGS of eight samples using *Leishmania* target enrichment (SureSelect Sequencing, SuSL-seq) was approved by the IRB of the Institute of Tropical Medicine, Antwerp (code 45/2024).

### Study site and Design

The study utilized 286 archived blood samples collected from patients with visceral leishmaniasis during the 2019-2022 VL outbreak in Garissa and Kitui counties, Kenya. Patients’ blood samples were collected at Garissa County referral hospital (S1 Fig.).

### DNA extraction and screening for *Leishmania* parasites

DNA was extracted from the archived blood samples using the MagMAX™ CORE, Nucleic Acid Purification reagents (Thermo Fisher Scientific, USA). For detection of *Leishmania* parasites, extracted DNA was screened at genus level by quantitative real-time PCR of the arginine permease gene AAP3 (AAP3-qRT-PCR). The following primer sets and probe were used [25]: forward - (AAP3), 5’ GGC GGC GGT ATT ATC TCG AT 3’, reverse - (AAP3), 5’ ACC ACG AGG TAG ATG ACA GAC A 3’ and probe - FAM 5’ ATGTCGGGCATCATC 3’ NFQ. All reactions were conducted in a 10.0μL reaction volume comprising 5.0µL of 2× SensiFAST master mix (Meridian Bioscience USA, https://www.meridianbioscience.com) (Meridian Bioscience, USA), 0.8µL of each primer at a concentration of 10 μM, 0.4µL of probe at concentration of 10 μM, 2.0 µL of DNA, and 1.0 µL of PCR water (Thermo Fisher Scientific, USA). Each reaction included known positive controls of *Leishmania* spp (*L. donovani* and *L. major* and non-target controls (PCR water) as negative controls to track contamination.

### Species identification and targeted genotyping

DNA was used to PCR amplify the *Hsp70* gene and ITS region using primer sets as published elsewhere [26] with minor modifications. The following primers were used: *Hsp70* gene forward (F25) 5’ -GGA CGC CAC GAT TKC T-3’ and reverse (R1310) 5’ -CCT GGT TGT TCA GCC ACT C-3’. For the ITS, the forward primer (LITSR): 5’ -CTG GAT CAT TTT CCG ATG-3’ and reverse primer (LITSV): 5’ -ACA CTC AGG TCT GTA AAC-3’ were used. The assays were conducted in 25μL reaction volume comprising: 12.5μL, 2X Mytaq Red Mix (Meridian Bioscience, USA), 0.5μL of each primer, 9.5μL of PCR grade water (Thermo Fisher Scientific, USA), and 2μL of the sample. The following conditions were used: For *Hsp70*: 94°C for 5 minutes, followed by 45 cycles of 94°C for 30 seconds, 61°C for 1 minute and 72°C for 3 minutes, followed by a final extension step at 72°C for 10 minutes. For ITS: 95°C for 2 minutes followed by 34 cycles of 95°C for 20 sec, 53°C for 30 sec, 72°C for 1 min, followed extension at 72°C for 6 min. The amplified products were visualized on a 2% agarose gel stained with 3.0μL GelRed™ (Biotium Inc., USA). Amplicons were cleaned using AmpureXP beads (Beckman Coulter, USA) following manufacturer’s instructions, and then used to construct sequencing libraries with the Collibri™ ES DNA Library Prep Kit from Illumina™ (Thermo Fisher scientific, USA). The individual libraries for both *Hsp70* and ITS loci were normalized to 4nM concentration and then pooled separately. The pooled amplicon library was denatured and diluted to a final concentration of 8pM, then spiked with 5% PhiX (Illumina, USA) as a sequencing control and sequenced on the MiSeq platform, with 600 v.3 paired end chemistry (Illumina, CA, USA).

### Whole genome sequencing by target enrichment

Eight samples with high parasitemia as measured by genus-specific PCR (Ct values between 26-33) were selected for WGS. *Leishmania* DNA quantification was then performed by qPCR as described by Domagalska et al. [21]. Starting concentration of DNA samples was between 3.04-9.09 ng/µl.

We used 21-50ng of total DNA for library preparation. DNA fragmentation was performed enzymatically using the Agilent SureSelect Enzymatic Fragmentation Kit (Agilent, Santa Clara, USA). Libraries were prepared using the SureSelect XT HS Target Enrichment System for Illumina (Agilent, Santa Clara, USA). Subsequently, adaptor-ligated libraries were prepared using 11 cycles in pre-capture PCR. Libraries were hybridized with the custom probes (Agilent, *L_donovani_infantum*, SuSL design ID S3377046, containing 318,358 probes, each 120 bp RNA oligonucleotides and covering 29.987 Mbp.) at a dilution 1:10, and captured with Dynabeads MyOne Streptavidin T1 magnetic beads (Thermo Fisher Scientific, Waltham, USA). After washing steps, the DNA captured by streptavidin beads was amplified by PCR and purified with AMPure XP beads. The quantity and quality of the libraries were assessed by TapeStation using High Sensitivity D1000 ScreenTape (Agilent Technologies, Santa Clara, USA). Sequencing was conducted on the Illumina NovaSeq platform using 2×150 bp paired reads at GenomeScan (Netherlands), for which 59,22-78,49 million raw reads were obtained (S3 Table).

### Analysis of Data (targeted sequencing)

AAP3-qRT-PCR results for positive samples at genus were tabulated as either negative or positive based on the set cycle threshold (Ct) values. For the *Hsp70* and ITS sequences, demultiplexed sequence reads from the Miseq were processed using the ngs_mapper (https://github.com/VDBWRAIR/ngs_mapper) pipeline which performed read trimming, reference mapping and consensus sequence generation. The consensus sequences were checked manually to resolve errors using Geneious prime 2022.2.1 (GraphPad LLC, MA, USA) and https://igv.org. Blastn v2.15.0 was used for homology analysis of the *Hsp70* (n=86) and ITS (n=79) sequences. Reference sequences obtained from GenBank (NC_018255.1 for *Hsp70* gene and NC_018254 for ITS region) were used to guide the homology analysis. To establish the phylogenetic relationship between Kenyan *Leishmania* spp and those from other regions, a comprehensive dataset of curated, annotated, and published *Leishmania* sequences were retrieved from NCBI GenBank (*Hsp70*, n=67 and ITS, n=65) (Supplementary Table 1) and aligned using the MAFFT plugin in Geneious prime v.2022.2.1 (Auckland, New Zealand).

The *Hsp70* and ITS sequences were first analyzed separately and then concatenated to improve the phylogenetic resolution of individual genes. The concatenated sequences (n=64) were aligned with previously published *Leishmania* sequences (n=36) (Supplementary Table 1). The individual and concatenated genes were used to infer Maximum-likelihood (ML) trees using the best fitting nucleotide substitution model as determined by ModelFinder [27] within IQ -tree v2.2.26 (https://pubmed.ncbi.nlm.nih.gov/28481363/). Branch support was determined using both the ultrafast bootstrap approximation (UFboot) with 1,000 replicates and the SH-like approximate likelihood ratio test (SH-aLRT) also with 1,000 bootstrap replicates. Visualization and annotation of the resulting phylogenetic trees was done using Figtree version 1.4.2 (http://tree.bio.ed.ac.uk/software/Figtree).

### Analysis of Data (whole genome sequencing)

For WGS by target enrichment, demultiplexed paired end reads from the NovaSeq6000 platform were assessed for quality using FastQC v0.14.1 (https://www.bioinformatics.babraham.ac.uk/projects/fastqc/) and FastQ Screen (https://www.bioinformatics.babraham.ac.uk/projects/fastq_screen/). In addition to the genomes sequenced in the context of this work, we also added a set of representatives of the different clusters as defined in Franssen et al. [28] as available under the NCBI BioProject number PRJEB2600, PRJEB2724, PRJEB8947 and PRJEB2115 (S1 Table).

Reads were aligned to the *L. donovani* BPK282 reference genome [29] using BWA (v0.7.17) [30] with a seed length of 50. Only properly paired reads with a mapping quality >30 were retained using SAMtools [31]. Duplicate reads were removed using Picard (v2.22.4) with the RemoveDuplicates function. SNP calling followed the GATK (v4.1.4.1) best practices workflow: (i) HaplotypeCaller was used to generate GVCF files for each sample; (ii) individual GVCFs were merged using CombineGVCFs; (iii) genotyping was performed with GenotypeGVCFs; and (iv) SNP and INDEL filtering was applied using SelectVariants and VariantFiltration, with filtering criteria based on GATK best practices (QD > 2, QUAL > 30, SOR < 3, FS < 60, MQ > 40, MQRankSum > -12.5, ReadPosRankSum > -8.0) [32]. Variant regions associated with known drug resistance markers were extracted using BCFtools [33]. Impact of those mutations was assess using the SnpEff software [34], and visualized with the pheatmap package in R. Overlap of SNPs between two strains was determined using the vcfR [35] (v1.13.0) and adegenet (v2.1.8) packages [36], with visualization performed using pheatmap (v1.0.12).

For phylogenetic analysis, bi-allelic SNPs were extracted from VCF files using BCFtools and converted to Phylip format with the vcf2phylip.py script (https://github.com/edgardomortiz/vcf2phylip). Phylogenetic trees were inferred using RAxML [37] under the GTR+G model, using *L. aethiopica* L100 as an outgroup. Tree visualization was performed with ggtree (v 3.14) [38]. To construct unrooted phylogenetic networks, bi-allelic SNPs were converted to FASTA format using the vcf2fasta.py script (https://github.com/FreBio/mytools/tree/master) and analyzed in SplitsTree4 using the NeighborNet algorithm [39].

For downstream population genomic analysis, SNP pruning for linkage disequilibrium was performed using PLINK 2.0 (--indep-pairwise 50 10 0.5) [40], followed by bed file generation. ADMIXTURE [41] was used to assess ancestry in both WGS and SureSelect samples, excluding clonal individuals within each phylogenetic group. The number of populations (K) was evaluated from 4 to 11 using a five-fold cross-validation procedure. Population structure inferred by ADMIXTURE was visualized with ggplot2 in R.

For the exploration of possible genomic signatures of drug resistance, we selected 11 loci that were previously shown to be involved in *L. donovani* resistance to drugs used in clinical practice. Relevant regions (indicated below) were subset using BCFtools [33]. Only SNPs having an effect at the protein level (missense and non-sense mutations) were retained. Visualization of the heatmap was performed using the pheatmap function in R.

## Results

### Screening of *Leishmania* parasites

The demographics associated with the *Leishmania* samples is shown in Table 1. Out of the 286 blood samples tested by AAP3-qRT-PCR, 128 (45%) were positive for *Leishmania* at the genus level. There was a 3:1 male:female infection ratio probably because of gender roles in this nomadic community. Men spend prolonged periods herding livestock where sand flies thrive. Additionally, seasonal migrations with livestock further expose them to diverse sand fly habitats, while women, who primarily engage in household activities, encounter less risk [13].

**Table 1.**
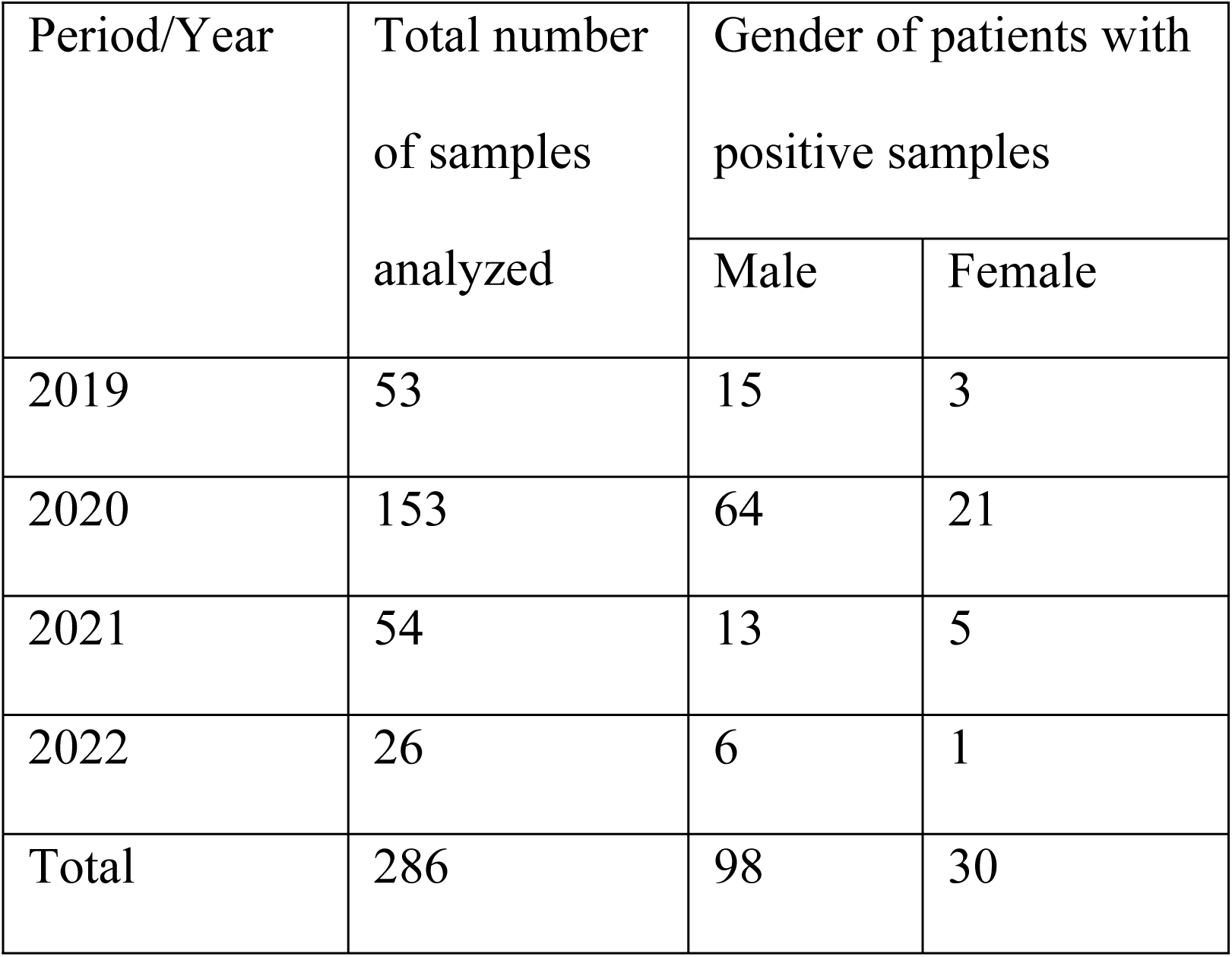
Patient demographics, all over the VL epidemics, from 2019 to 2022.

### Phylogenetic trees from individual genes

After quality control and assembly, we obtained consensus sequences of sizes 1,292 bp for *Hsp70* and 1,060 bp for ITS loci. Of the 128 samples positive for *Leishmania*, 86 generated usable sequences for *Hsp7*0 gene and 79 for the ITS region. Homology analysis of the *Hsp70* (S2 Fig.) and ITS (S3 Fig.) sequences revealed monophyletic grouping of study samples that clustered with the *L. donovani* species complex obtained from the NCBI GenBank.

The closest relatives of the study sequences inferred from individual genes differed, with *Hsp70* (S2 Fig.) showing phylogenetic proximity to countries north of Kenya (Sudan, Ethiopia) and ITS (S3 Fig.) showing phylogenetic proximity to India, France, Spain, Morocco, Sri-Lanka, Sudan and Ethiopia. Given the different phylogenetic histories of *Hsp70* and ITS genes, we concatenated 64 study genomes against similar genomes available from the GenBank. In the concatenated tree, the study genomes clustered with the *L. donovani* complex in a group with *L. donovani* BPK282A1 (Fig. 1). Like in S3 Fig., the study sample Kenya/GSA-164/2020 remained an outlier in the concatenated tree (Fig. 1, shown in red star) and branched with the main *L. major* cluster, in a well-supported branch (bootstrap support of 63%). In the ITS tree, this sample, shown in red star (S3 Fig.) separated distinctly from the other study samples and formed a distinct outgroup. In the *Hsp70* tree, this genome was not distinct from the other study genomes.

**Fig. 1.**
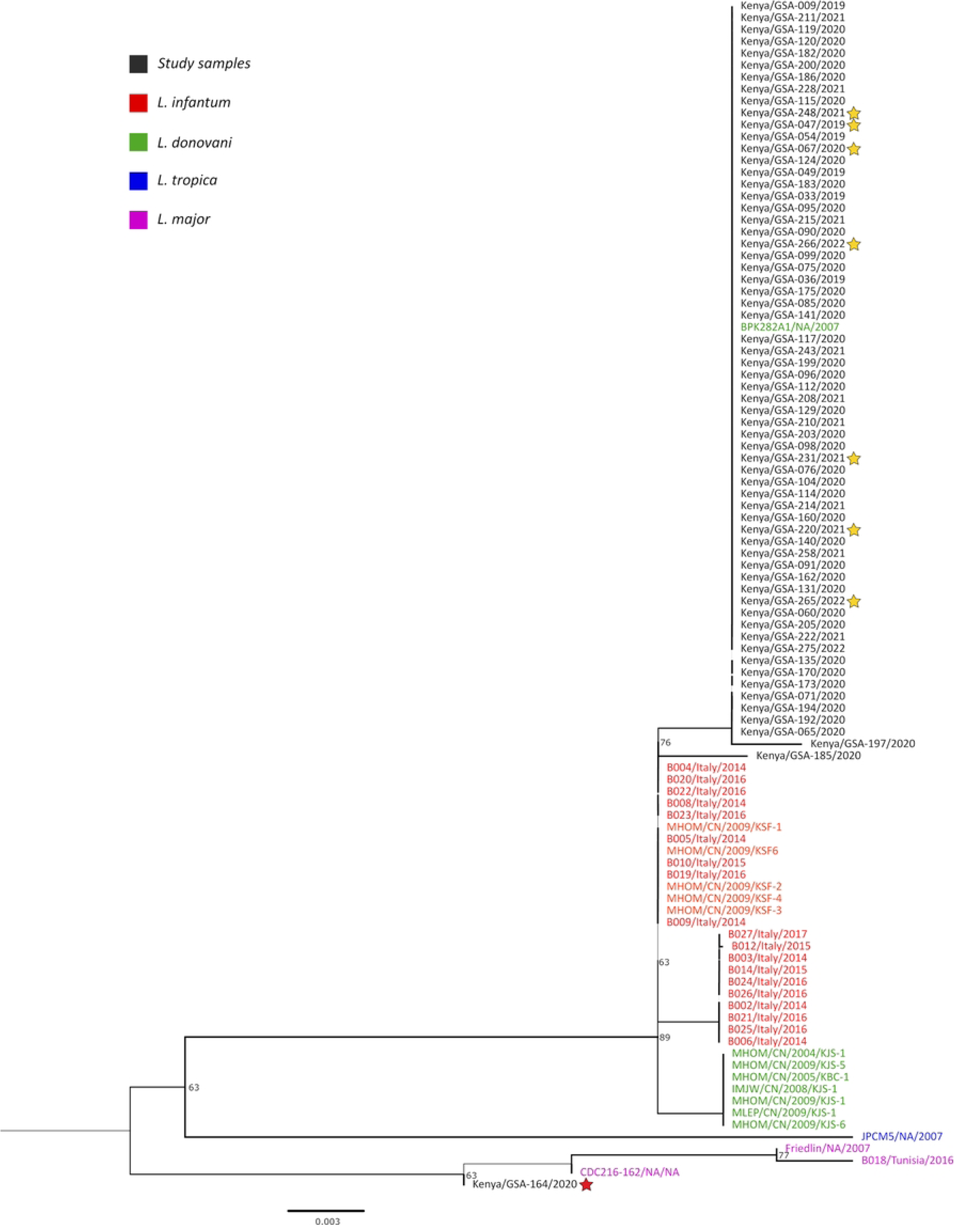
Maximum-likelihood phylogenetic tree inferred from an alignment of concatenated study *Hsp70* and ITS sequences (black) and select *Leishmania* sequences retrieved from NCBI GenBank (colored text). Red star shows an outlier study sample (Kenya/GSA-164/2020) that branched with the main *L. major* cluster. The yellow stars shows eight of the current study samples that were selected for targeted whole genome sequencing (see Fig. 2). Branch support was estimated using SH-like likelihood ratio test and are indicated as numbers. The scale bar represents the number of substitutions per site.

### Direct *Leishmania* genome sequencing in clinical samples after target enrichment

Based on parasite load, eight blood samples that Ct values of <33 were selected (S2 Table) for direct targeted sequencing by SuSL-seq. Summary of sequencing performances can be found in S3 Table. Phylogenetic analysis of the eight genomes together with published genomes of *L. donovani* (including groups Ld1-5, following classification by Franssen et al. [28], *L. infantum* and the three dermotropic species (*L. tropica, L. major* and *L. aethiopica*) [28] allowed us to identify the *Leishmania* species in these 8 Kenyan samples. They clearly branched in the *L. donovani* s.s. cluster, separately from *L. infantum* and the dermotropic species (Fig. 2). The eight Kenyan samples formed two distinct clades. Five of these samples clustered within the Ld5 group, which includes isolates from Kenya and Ethiopia collected between 1954 (LRC-L53) and 2009 (AM560WTI) [28]. The remaining three samples clustered together close to three ’other_Ldon’ samples (LRV-L740 from Israel, GE, and LEM3472 from Sudan) in a distinct intermediate position in the rooted phylogenetic tree, between the Ld3 group (Sudan/Ethiopia) and Ld4 group (Iraq) (Fig. 2). In the reticulated network, the three K-Ldnew samples branched separately from ‘other Ldon’ samples (S4 Fig.), suggesting an independent evolutionary origin. Interestingly, both Kenya’s Ld5 (K-Ld5) and the intermediate Kenyan samples (K-Ldnew) were present in both Garissa (Balambala and Kitui (Mwingi North) counties (S1 Fig.).

**Fig. 2.**
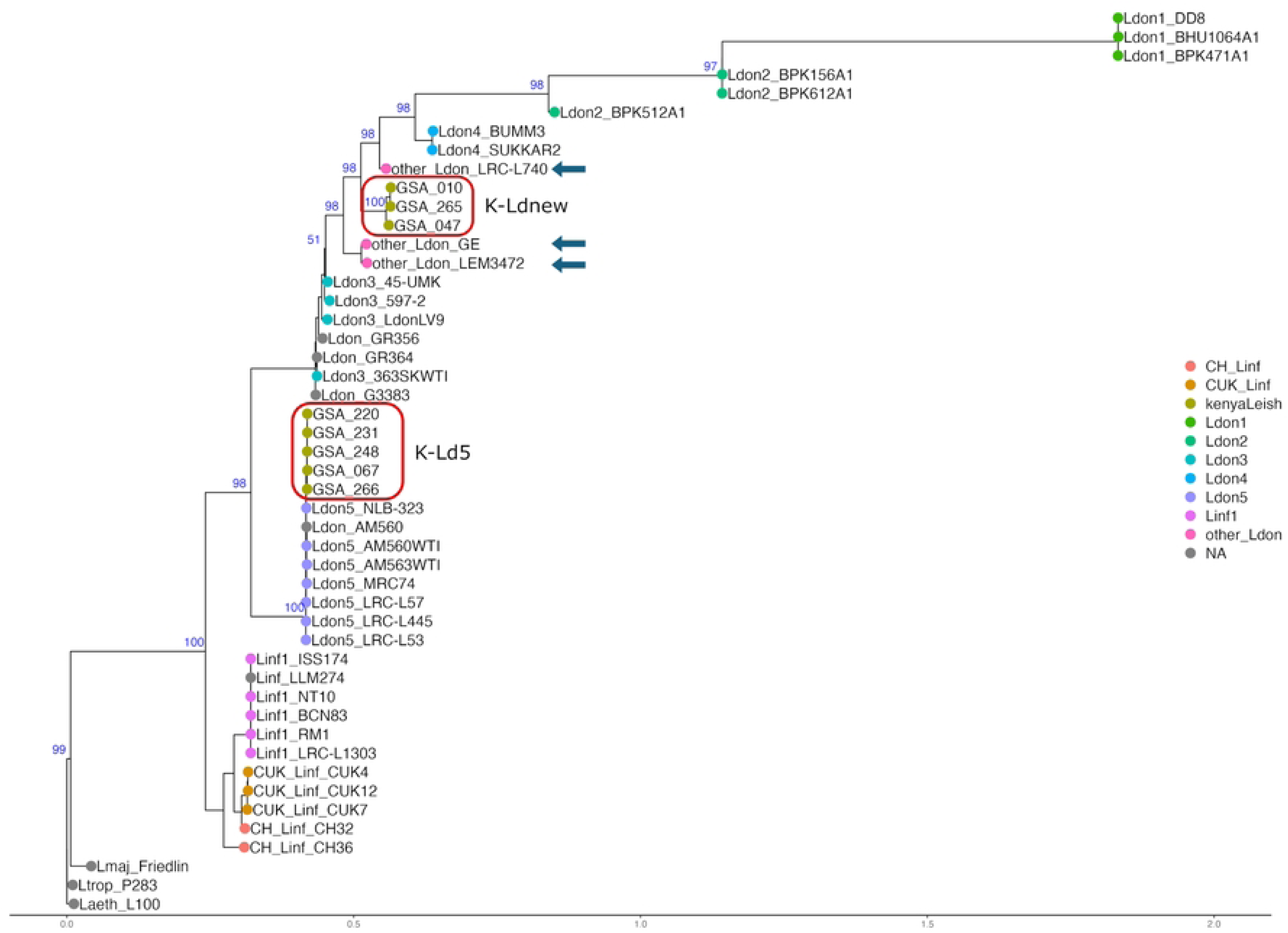
Rooted phylogenetic tree based on genome-wide bi-allelic SNPs inferred using RAxML, with *L. aethiopica* L147 as the outgroup (not shown). Most relevant bootstrap values are indicated in blue. The color code represents clustering as proposed by Franssen et al. [28] with samples from this study highlighted in olive green (“kenyaLeish”). The latter samples are distributed across two clusters, which are indicated by a red box. The three arrows indicate ‘Other-Ldon’ hybrid genotypes identified by Franssen et al. [28].

Given the intermediate phylogenetic position of the three Kenyan samples, we assessed if this genotype could have been generated by recombination between the previously reported *L. donovani* groups. Analysis of population structure was done in ADMIXTURE for *L. donovani* based on 61,186 genome-wide bi-allelic SNPs. When testing different number of populations in the admixture analyses, line plot shows the lowest cross-validation error (CV) between 4 to 7 populations (S5 Fig.). Mixed ancestry (Ld3 and Ld4) is shown for isolates LRC-L740, LEM3472 and GE as reported earlier [28] and the same pattern was observed in the three K-Ldnew samples (Fig.3). In contrast, K-Ld5 samples showed a single ancestral origin. Presence of mixed ancestry signatures in a sample could be reflecting events of genetic exchange as well as polyclonality (mixture of different genotypes). By plotting the alternative allele frequency along the 36 chromosomes of admixed sample GSA047, we found the signatures of genetic exchange, i.e. alternation of stretches of heterozygosity and homozygosity and average allele frequency of 50/50 in heterozygous stretches (see S6 Fig.)

**Fig. 3.**
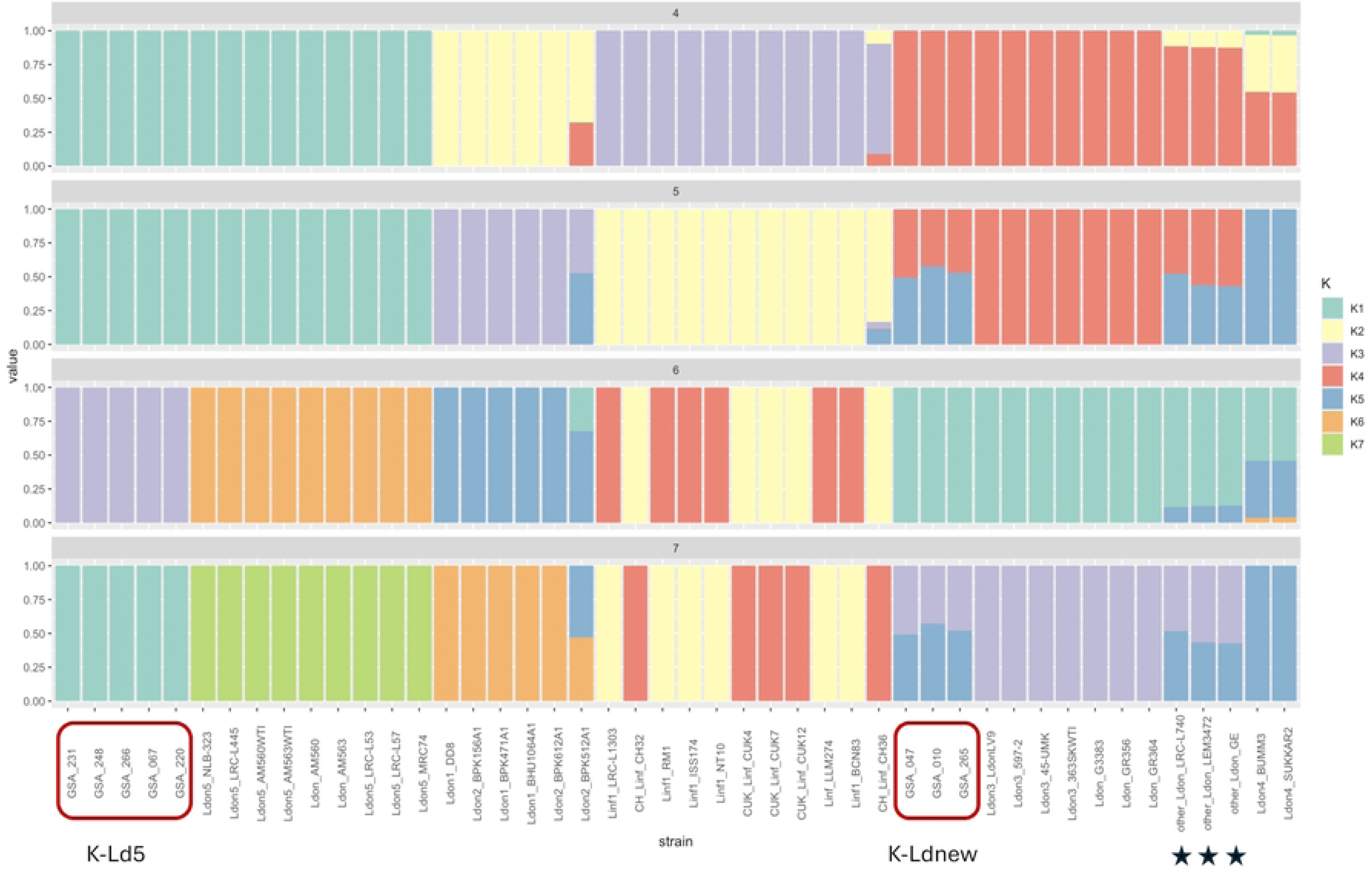
Admixture analysis (K between 4 and 7, see S5 Fig.); the red boxes correspond to the 8 samples from Kenya (this study); each bar represents a sample, with colors corresponding to ancestry components. the stars indicate the genotypes with mixed ancestry reported by Franssen et al. [28].

Finally, a scan of genomic signatures of resistance was undertaken on K-Ld5, the three K-Ldnew samples and a series of reference sequences. We first compiled a list of 11 genes reported to be involved in resistance to Antimony (Sb), Amphotericine B (AmB) and Miltefosine (MIL) [42–46] and analyzed SNPs having an effect at the protein level (missense and non-sense mutations). Overall, 46 SNPs were identified in the different genes that were analyzed. Of the 46 SNPs, 16 homozygous mutations related with resistance to Sb (MRPA), AmB (C5D) and MIL (MSL) were shared among the eight Kenyan samples and all Ld5 reference genomes and LRC-L740 (Fig.4). LiMT, the transporter of Miltefosine showed mutations in all Kenyan samples, but patterns differed between K-Ld5 and the intermediate Kenyan samples: respectively, positions 486 and 183 and positions 248 and 495. In addition, one of the K-Ldnew samples showed a mutation in AQP1 gene.

**Fig. 4.**
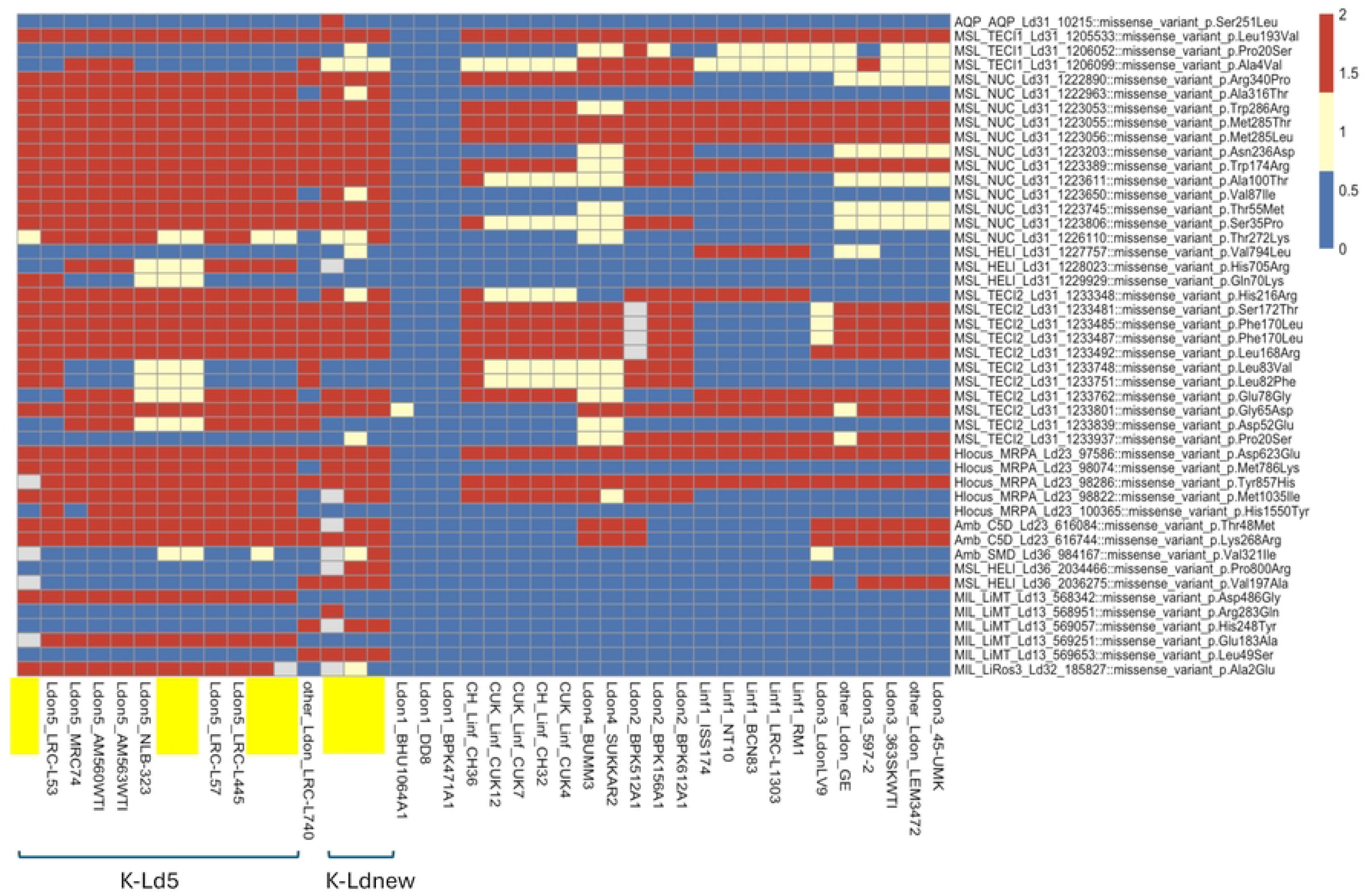
Heatmap showing the distribution of single nucleotide polymorphisms (SNPs) in 11 genes reported to be associated with drug-resistant phenotypes. The color scheme represents different SNP categories: blue indicates the absence of SNPs, orange indicates heterozygous SNPs, and red indicates homozygous SNPs. The naming convention for SNPs follows the format of the gene of interest, position in the genome, type of mutation, and its effect on the corresponding protein.

## Discussion

This study, for the first time in Kenya, used targeted and whole genome sequences from blood samples to characterize genomes of *Leishmania* parasites collected during the 2019-2022 VL outbreak in Garissa county. Multiple studies have documented outbreaks of VL in arid areas of North Eastern Kenya (Marsabit, Isiolo and Wajir counties) [47] and identified the influence of various environmental, ecological and social factors such as the presence of termite mounds around homesteads, animal burrows, residing in rural sub-counties, and being male as being associated with higher VL infection rates [48].

In the current study, *Leishmania* genomes were first analyzed by targeting the *Hsp70* and ITS loci. *Hsp70* is a conserved housekeeping gene that is essential for the parasite’s survival and adaptation to various environmental stresses and has been widely used for phylogeny and taxonomy of *Leishmania* parasites [49,50]. ITS region is extensively variable and is widely used for identification as well as phylogenetic analysis of *Leishmania* spp. [51].

Phylogenetically, all the 86 *Hsp70* sequences mapped to *L. donovani* complex from East African regions, indicating geographical kinship as has been suggested before [52]. Another interesting observation is the monophyletic nature of the study samples and those obtained from the NCBI GenBank, indicating the slow rate of evolution of the *L. donovani Hsp70* gene. For, instance we observed a Kenyan sample collected in 1955 (S2 Fig., black star) clustering with our current study samples. We also noted that all samples from our study clustered closely with those from Ethiopia (S2 Fig.).

Similar to *Hsp70*, the ITS phylogenetic tree was monophyletic (S3 Fig.). But, unlike the *Hsp70*, the phylogenetic proximity of the study samples inferred using the ITS was India, France, Spain, Morocco, Sri-Lanka, Sudan and Ethiopia. This discrepancy arises from the fact that the *Hsp70* and ITS sequences deposited in the GenBank do not necessarily come from the same regions. Interestingly, one sample (Kenya/GSA_164/2020) separated distinctly from the other samples (S3 Fig., red star). By *Hsp70*, this sample did not resolve differently from the other study genomes. To improve the resolution and accuracy of the phylogenetic trees generated from individual genes, the *Hsp70* and ITS data were concatenated into single data set (Fig. 1). In the concatenated tree, the study genomes clustered with the *L. donovani* complex. Like in the ITS data (S3 Fig.), the study sample Kenya/GSA-164/2020 remained an outlier and branched with the main *L. major* cluster, in a well-supported branch (bootstrap support of 63%).

We also combined target enrichment and WGS (SuSL-seq) for a detailed exploration of the *Leishmania* genome in 8 of the study samples. As shown in Fig.2, two distinct groups were identified: Group one comprising 5 study genomes that is related to *L. donovani* group 5 (Ld5) and referred here as K-Ld5 (K=Kenya). Group two comprised 3 samples of mixed ancestry, here referred to as K-LdNew. The SuSL-seq protocol/application was developed to study parasite genomes directly from their natural environment and to avoid the strong selection biases associated with WGS of cultivated parasite isolates [21]. Indeed, in a previous study that used *Leishmania* samples from Nepal, it was demonstrated that the genome of parasites directly sequenced in bone marrow of VL patients was systematically different from the genome of in vitro derived isolates [21]. This led to recommendation that, studies needing to establish a link between parasite genome and clinical or epidemiological features should use parasites sequenced directly from clinical samples [53]. This led to the use of SuSL-seq for source tracing of *L. donovani* in emerging foci of VL in West of Nepal [22]. Direct sequencing of *Leishmania* parasites from blood has major advantages, among them facilitating sampling in remote places with minimal laboratory facilities and allows discovery of new genotypes such as ISC11 described in the Nepal study [21] or the intermediate Kenyan genotypes (K-Ld5-New) in the present study. It is currently unknown whether these seemingly new genotypes represent new genotypes or were under-reported because they were less adapted to culture conditions or represent true emergence of new genotypes in the study region. More direct sequencing studies involving past collections of *Leishmania* samples are needed in order to rule out sampling bias.

Given the phylogenetic proximity of the K-Ldnew parasites and isolates LRV-L740, GE and LEM3472 in the phylogenetic tree (Fig.2), together with the admixture pattern similarity between these samples (Supp. Fig. 5b), we propose to call this group Ld6, hereby complementing the Ld1-Ld5 classification proposed by Franssen et al. [28]. Noteworthy, this group is not monophyletic and is a matter of convenience to class together parasites resulting from different admixture events between well differentiated populations Ld3 and Ld4. as shown by the reticulated network (S4 Fig.) and previously hypothesized for LRV-L740, GE and LEM3472 [28].

Our study illustrates the genetic diversity within *L. donovani* complex, especially in East Africa where VL has become a hot spot. The observation of intra-specific genetic admixture confirms a trend increasingly reported in several species all over the world [54,55] and is likely explained by the high discriminatory power of genome sequencing approaches. In addition, in the selected genes reported to be involved in drug resistance, we identified a total of 46 SNPS in all the 8 Kenyan samples, 16 of which have been found to code for resistance to Antimony, Amphotericine B and Miltefosine [42–46]. Nevertheless, the presence of these drug resistance genes should be interpreted with care: at this stage, the genes represent genomic signatures and further work is required to understand their functional implication in resistance. Further research is needed to extend our sequencing approach to other regions in Kenya and surrounding countries and assess among others the epidemiological dynamics and the dispersal/spread of parasites across the continent and between continents (for instance Africa and the Indian sub-continent). Present studies and previous ones make a strong case for genomic surveillance studies using clinical samples so as to understand the changing landscape of *L. donovani*, which is essential to support control programs.

## Data availability

All sequencing data are available at http://www.ncbi.nlm.nih.gov/bioproject/1251555 (PRJNA1251555) for whole genome sequences. *Hsp70* and ITS loci are available under GenBank accession numbers PQ700286–PQ700371 and PV557379–PV557457, respectively. All other data are available in the main text or supplementary material.

## Acknowledgments.

We are grateful to the technical field assistants who supported the sample collection and logistics in Garissa County Referral Hospital. We also thank the patients who participated in this study. Special thanks to Rachael Githii for generating the map shown in S1 Fig.

This study was supported by the Armed Forces Health Surveillance Division and its Global Emerging Infections Surveillance and Research Branch (grant numbers ProMIS P0089_24_KY). SH was supported by the Research Foundation Flanders (grants G092921N); MAD acknowledges support from Flemish Ministry of Science and Innovation.

## Supporting Information

**S1 Fig.** Map of Garissa and Kitui counties, Kenya, indicating the location of Garissa County Referral Hospital, where the visceral leishmaniasis samples were collected. The dots indicate the reported origin of the 8 *Leishmania* samples (3 in Garissa and 5 in Kitui) analyzed by whole genome target (WGS) enrichment (SureSelect Sequencing, SuSL-seq). K-Ld5 and K-Ldnew correspond to two genomic groups identified after WGS analysis (see Fig.2). Genomic group one refers to *L. donovani* group 5 (K-Ld5: K for Kenya) similar to the previously reported *L. donovani* group 5. The other constituted a new genetic variant not reported previously (K-Ldnew).

**S1 Table.** Genomes used in this study

**S2 Fig**. Maximum-likelihood phylogenetic tree showing the relationship between *Hsp70* gene sequences of the study samples (black) and selected *Leishmania* sequences retrieved from NCBI GenBank (colored fonts). Black star shows a *L. donovani* sequence from Kenya that was deposited in GenBank in 1955 clustering with current study samples. The yellow stars shows 8 of the current study samples that were selected for targeted whole genome sequencing (see Figure 3). Branch support was estimated using SH-like likelihood ratio test and is shown as thick lines where branch support > 50%. The scale bar represents the number of substitutions per site.

**S3 Fig.** Maximum-likelihood phylogenetic tree showing the relationship between the ITS sequences of the study (black) and selected *Leishmania* sequences retrieved from NCBI GenBank (colored fonts). Red star shows an outlier study sample (Kenya/GSA-164/2020), separating distinctly from others. The yellow stars shows 8 of the current study samples that were selected for targeted whole genome sequencing (see Fig. 3). Branch support was estimated using SH-like likelihood ratio test and is shown as thick lines where branch support > 50%. The scale bar represents the number of substitutions per site.

**S2 Table**. Demographics for eight blood samples selected for targeted whole genome sequencing.

**S3 Table.** Summary of sequencing performances

**S4 Fig.** Phylogenetic network created based on all bi-allelic SNPs using the NeighborNet algorithm as implemented in SplitsTree4. The samples in this study overlapping with cluster 5 *L. donovani* strains – as defined in Franssen et al., (2020) are indicated with a green ellipse. The three other strains not grouping with any of the known clusters are indicated in a red ellipse.

**S5 Fig.** ADMIXTURE analysis. Cross validation error as extracted from the ADMIXTURE log files for each tested value of the number of populations (K).

**S6 Fig.** Allele frequency plot for 12 chromosomes in two samples, admixed (K-Ldnew GSA047) and with single ancestry origin (K-Ld5, GSA248). The X-axis represents the position of the SNP in the chromosome, and the Y-axis represents the allele frequency of the SNP in the corresponding strain. Presence of mixed ancestry signatures in a sample could be reflecting events of genetic exchange as well as polyclonality (mixture of different genotypes). To distinguish the two hypotheses, we plotted the alternative allele frequencies along the 36 chromosomes of admixed samples. In case of polyclonal mixture, alternative allele frequency should be constant all along the chromosomes and it should deviate from 50/50. While in case of genetic exchange, stretches of heterozygosity should be alternating with stretches of homozygosity (intra-chromosomal recombination) and in heterozygous stretches, the frequency should always be 50/50. The latter was observed for most chromosomes of admixed sample, not for the sample with single ancestry origin. Noteworthy, the analysis could only be done with samples with similar and high genome coverage (see S3 Table): samples GSA47 and GSA248 show a coverage > 10% in 65,5 and 85.6% of the genome respectively (the two other admixed samples GSA010 and GSA265 showed only 4.4 and 26.5% of genome coverage).

## Notes

### Competing Interest Statement

The authors have declared no competing interest.

### Funding Statement

Yes

### Author Declarations

This study was conducted as part of a public health response following a request from the Ministry of Health, Garissa County to support a Leishmania outbreak investigation. The WRAIR Human Subject Protection Branch determined that the outbreak response did not meet the definition of research and does not require IRB review. WGS of eight samples using Leishmania target enrichment (SureSelect Sequencing, SuSL-seq) was approved by the IRB of the Institute of Tropical Medicine, Antwerp (code 45/2024).

## References

1. Reithinger R, Dujardin J-C, Louzir H, Pirmez C, Alexander B, Brooker S. Cutaneous leishmaniasis. Lancet Infect Dis. 2007;7: 581–596. doi:10.1016/S1473-3099(07)70209-8

2. WHO. Leishmaniasis: The Global Health Observatory [Internet]. 2023. Available: https://www.who.int/data/gho/data/themes/topics/gho-ntd-leishmaniasis

3. Akuffo H, Costa C, van Griensven J, Burza S, Moreno J, Herrero M. New insights into leishmaniasis in the immunosuppressed. PLoS Negl Trop Dis. 2018;12: e0006375.

4. McCall L-I, Zhang W-W, Matlashewski G. Determinants for the Development of Visceral Leishmaniasis Disease. PLoS Pathog. 2013;9: e1003053.

5. Pace D. Leishmaniasis. J Infect. 2014;69 Suppl 1: S10-8. doi:10.1016/j.jinf.2014.07.016

6. Rojas-Jaimes J, Rojas-Palomino N, Pence J, Lescano AG. Leishmania species in biopsies of patients with different clinical manifestations identified by high resolution melting and nested PCR in an Endemic district in Peru. Parasite Epidemiol Control. 2019;4: e00095. 10.1016/j.parepi.2019.e00095

7. Wamai RG, Kahn J, McGloin J, Ziaggi G. Visceral leishmaniasis: a global overview. J Glob Health Sci. 2020;2.

8. Dahl EH, Hamdan HM, Mabrouk L, Matendechero SH, Mengistie TB, Elhag MS, et al. Control of visceral leishmaniasis in East Africa: fragile progress, new threats. BMJ Glob Health. 2021;6: e006835. doi:10.1136/bmjgh-2021-006835

9. WHO. Eliminating visceral leishmaniasis: India takes decisive steps to overcome last-mile challenges. 2020. Available: https://www.who.int/news/item/05-03-2020-VL-India-takes-decisive-steps-overcome-last-mile-challenges

10. Pandey DK, Alvar J, den Boer M, Jain S, Gill N, Argaw D, et al. Kala-azar elimination in India: reflections on success and sustainability. Int Health. 2025; ihaf013. doi:10.1093/inthealth/ihaf013

11. Jones CM, Welburn SC. Leishmaniasis Beyond East Africa. Front Vet Sci. 2021;8: 618766. doi:10.3389/fvets.2021.618766

12. Ouma FF, Mulambalah CS. Persistence and Changing Distribution of Leishmaniases in Kenya Require a Paradigm Shift. J Parasitol Res. 2021;2021: 9989581. doi:10.1155/2021/9989581

13. Kanyina EW. Characterization of visceral leishmaniasis outbreak, Marsabit County, Kenya, 2014. BMC Public Health. 2020;20: 446. doi:10.1186/s12889-020-08532-9

14. Akhoundi M, Kuhls K, Cannet A, Votýpka J, Marty P, Delaunay P, et al. A Historical Overview of the Classification, Evolution, and Dispersion of Leishmania Parasites and Sandflies. PLoS Negl Trop Dis. 2016;10: e0004349. doi:10.1371/journal.pntd.0004349

15. Kevric I, Cappel MA, Keeling JH. New world and old world Leishmania infections: a practical review. Dermatol Clin. 2015;33: 579–593.

16. Mitropoulos P, Konidas P, Durkin-Konidas M. New World cutaneous leishmaniasis: updated review of current and future diagnosis and treatment. J Am Acad Dermatol. 2010;63: 309–322.

17. Lau R, Mukkala AN, Kariyawasam R, Clarke S, Valencia BM, Llanos-Cuentas A, et al. Comparison of Whole Genome Sequencing versus Standard Molecular Diagnostics for Species Identification in the Leishmania Viannia Subgenus. Am J Trop Med Hyg. 2021;105: 660–669. doi:10.4269/ajtmh.21-0273

18. Tonui WK, Titus RG. Leishmania major soluble exo-antigens (Lm SEAgs) protect neonatal BALB/c mice from a subsequent challenge with L. major and stimulate cytokine production by Leishmania-naïve human peripheral blood mononuclear cells. Journal of Parasitology. 2006;92: 971–976.

19. Akhoundi M, Downing T, Votýpka J, Kuhls K, Lukeš J, Cannet A, et al. Leishmania infections: Molecular targets and diagnosis. Mol Aspects Med. 2017;57: 1–29. 10.1016/j.mam.2016.11.012

20. Desjeux P. The increase in risk factors for leishmaniasis worldwide. Trans R Soc Trop Med Hyg. 2001;95: 239–243.

21. Domagalska MA, Imamura H, Sanders M, Van den Broeck F, Bhattarai NR, Vanaerschot M, et al. Genomes of Leishmania parasites directly sequenced from patients with visceral leishmaniasis in the Indian subcontinent. PLoS Negl Trop Dis. 2019;13: 1–22. doi:10.1371/JOURNAL.PNTD.0007900

22. Monsieurs P, Cloots K, Uranw S, Banjara MR, Ghimire P, Burza S, et al. Source Tracing of Leishmania donovani in Emerging Foci of Visceral Leishmaniasis, Western Nepal. Emerg Infect Dis. 2024;30: 611.

23. Hassaballa IB, Torto B, Sole CL, Tchouassi DP. Exploring the influence of different habitats and their volatile chemistry in modulating sand fly population structure in a leishmaniasis endemic foci, Kenya. PLoS Negl Trop Dis. 2021;15: e0009062.

24. Boussery G, Boelaert M, Van Peteghem J, Ejikon P, Henckaerts K. Visceral Leishmaniasis (Kala-Azar) Outbreak in Somali Refugees and Kenyan Shepherds, Kenya. Emerging Infectious Disease journal. 2001;7: 603. doi:10.3201/eid0707.017746

25. Tellevik MG, Muller KE, Løkken KR, Nerland AH. Detection of a broad range of Leishmania species and determination of parasite load of infected mouse by real-time PCR targeting the arginine permease gene AAP3. Acta Trop. 2014;137: 99–104. 10.1016/j.actatropica.2014.05.008

26. Chen Y-F, Liao L-F, Wu N, Gao J-M, Zhang P, Wen Y-Z, et al. Species identification and phylogenetic analysis of Leishmania isolated from patients, vectors and hares in the Xinjiang Autonomous Region, The People’s Republic of China. PLoS Negl Trop Dis. 2021;15: e0010055.

27. Nguyen L-T, Schmidt HA, Von Haeseler A, Minh BQ. IQ-TREE: a fast and effective stochastic algorithm for estimating maximum-likelihood phylogenies. Mol Biol Evol. 2015;32: 268–274.

28. Franssen SU, Durrant C, Stark O, Moser B, Downing T, Imamura H, et al. Global genome diversity of the \textit{Leishmania donovani} complex. Weigel D, Clayton C, Ouellette M, editors. Elife. 2020;9: e51243. doi:10.7554/eLife.51243

29. Dumetz F, Imamura H, Sanders M, Seblova V, Myskova J, Pescher P, et al. Modulation of Aneuploidy in Leishmania donovani during Adaptation to Different In Vitro and In Vivo Environments and Its Impact on Gene Expression. mBio. 2017;8. doi:10.1128/mBio.00599-17

30. Li H, Durbin R. Fast and accurate short read alignment with Burrows–Wheeler transform. Bioinformatics. 2009;25: 1754–1760. doi:10.1093/bioinformatics/btp324

31. Li H, Handsaker B, Wysoker A, Fennell T, Ruan J, Homer N, et al. The Sequence Alignment/Map format and SAMtools. Bioinformatics. 2009;25: 2078–2079. doi:10.1093/bioinformatics/btp352

32. McKenna A, Hanna M, Banks E, Sivachenko A, Cibulskis K, Kernytsky A, et al. The Genome Analysis Toolkit: a MapReduce framework for analyzing next-generation DNA sequencing data. Genome Res. 2010;20: 1297–1303. doi:10.1101/gr.107524.110

33. Danecek P, Bonfield JK, Liddle J, Marshall J, Ohan V, Pollard MO, et al. Twelve years of SAMtools and BCFtools. Gigascience. 2021;10: giab008. doi:10.1093/gigascience/giab008

34. Cingolani P, Adrian P, Le Lily W, Melissa C, Tung N, Luan W, et al. A program for annotating and predicting the effects of single nucleotide polymorphisms, SnpEff. Fly (Austin). 2012;6: 80–92. doi:10.4161/fly.19695

35. Knaus B, Grünwald N. VCFR: a package to manipulate and visualize variant call format data in R. Mol Ecol Resour. 2016;17. doi:10.1111/1755-0998.12549

36. Jombart T, Ahmed I. adegenet 1.3-1: new tools for the analysis of genome-wide SNP data. Bioinformatics. 2011;27: 3070–3071. doi:10.1093/bioinformatics/btr521

37. Kozlov AM, Darriba D, Flouri T, Morel B, Stamatakis A. RAxML-NG: a fast, scalable and user-friendly tool for maximum likelihood phylogenetic inference. Bioinformatics. 2019;35: 4453– 4455. doi:10.1093/bioinformatics/btz305

38. Yu G. Using ggtree to Visualize Data on Tree-Like Structures. Curr Protoc Bioinformatics. 2020;69: e96. 10.1002/cpbi.96

39. Huson DH, Bryant D. Application of Phylogenetic Networks in Evolutionary Studies. Mol Biol Evol. 2006;23: 254–267. doi:10.1093/molbev/msj030

40. Chang CC, Chow CC, Tellier LCAM, Vattikuti S, Purcell SM, Lee JJ. Second-generation PLINK: rising to the challenge of larger and richer datasets. Gigascience. 2015;4: s13742-015-0047–8. doi:10.1186/s13742-015-0047-8

41. Alexander DH, Novembre J, Lange K. Fast model-based estimation of ancestry in unrelated individuals. Genome Res. 2009;19: 1655–1664. doi:10.1101/gr.094052.109

42. Potvin J-E, Leprohon P, Queffeulou M, Sundar S, Ouellette M. Mutations in an Aquaglyceroporin as a Proven Marker of Antimony Clinical Resistance in the Parasite Leishmania donovani. Clin Infect Dis. 2021;72: e526–e532. doi:10.1093/cid/ciaa1236

43. Dumetz F, Cuypers B, Imamura H, Zander D, D’Haenens E, Maes I, et al. Molecular Preadaptation to Antimony Resistance in Leishmania donovani on the Indian Subcontinent. mSphere. 2018;3. doi:10.1128/mSphere.00548-17

44. Pountain AW, Weidt SK, Regnault C, Bates PA, Donachie AM, Dickens NJ, et al. Genomic instability at the locus of sterol C24-methyltransferase promotes amphotericin B resistance in Leishmania parasites. PLoS Negl Trop Dis. 2019;13: e0007052. doi:10.1371/journal.pntd.0007052

45. Pérez-Victoria FJ, Sánchez-Cañete MP, Castanys S, Gamarro F. Phospholipid translocation and miltefosine potency require both L. donovani miltefosine transporter and the new protein LdRos3 in Leishmania parasites. J Biol Chem. 2006;281: 23766–23775. doi:10.1074/jbc.M605214200

46. Carnielli JBT, Dave A, Romano A, Forrester S, de Faria PR, Monti-Rocha R, et al. 3&#x2032;Nucleotidase/nuclease is required for *Leishmania infantum* clinical isolate susceptibility to miltefosine. EBioMedicine. 2022;86. doi:10.1016/j.ebiom.2022.104378

47. Organization WH. Leishmaniasis Fact sheet N 375. Geneva, Switzerland: WHO Media Centre. 2015.

48. Dulacha D, Mwatha S, Lomurukai P, Owiny MO, Matini W, Irura Z, et al. Epidemiological characteristics and factors associated with Visceral Leishmaniasis in Marsabit County, Northern Kenya. J Interv Epidemiol Public Heal. 2019;2: 3.

49. Fraga J, Montalvo AM, De Doncker S, Dujardin J-C, Van der Auwera G. Phylogeny of Leishmania species based on the heat-shock protein 70 gene. Infect Genet Evol. 2010;10: 238–245. doi:10.1016/j.meegid.2009.11.007

50. Jariyapan N, Bates MD, Bates PA. Molecular identification of two newly identified human pathogens causing leishmaniasis using PCR-based methods on the 3′ untranslated region of the heat shock protein 70 (type I) gene. PLoS Negl Trop Dis. 2021;15: e0009982.

51. Cupolillo E, Grimaldi Jr G, Momen H, Beverley SM. Intergenic region typing (IRT): a rapid molecular approach to the characterization and evolution of Leishmania. Mol Biochem Parasitol. 1995;73: 145–155.

52. Pigott DM, Bhatt S, Golding N, Duda KA, Battle KE, Brady OJ, et al. Global distribution maps of the leishmaniases. Elife. 2014;3: e02851.

53. Domagalska MA, Dujardin J-C. Next-generation molecular surveillance of TriTryp diseases. Trends Parasitol. 2020;36: 356–367.

54. Heeren S, Maes I, Sanders M, Lye L-F, Adaui V, Arevalo J, et al. Diversity and dissemination of viruses in pathogenic protozoa. Nat Commun. 2023;14: 8343. doi:10.1038/s41467-023-44085-2

55. Lypaczewski P, Matlashewski G. Leishmania donovani hybridisation and introgression in nature: a comparative genomic investigation. Lancet Microbe. 2021;2: e250–e258. 10.1016/S2666-5247(21)00028-8

